# Leveraging Illumina iSeq100 for Whole Genome Sequencing of *Salmonella Typhi*: a practical approach for resource-limited setting

**DOI:** 10.1101/2024.09.22.24314150

**Authors:** Nishan Katuwal, Shiva Ram Naga, Aastha Shrestha, Sabin Bikram Shahi, Dipesh Tamrakar, Rajeev Shrestha

**Affiliations:** Molecular and Genome Sequencing Research Lab, Dhulikhel Hospital, Kathmandu University Hospital, Dhulikhel, Nepal; Center for Infectious Disease Research and Surveillance, Dhulikhel Hospital, Kathmandu University Hospital, Dhulikhel, Nepal; Research and Development Division, Dhulikhel Hospital, Kathmandu University Hospital, Dhulikhel, Nepal; Department of Community Medicine, Kathmandu University School of Medical Sciences, Dhulikhel, Nepal; Department of Pharmacology, Kathmandu University School of Medical Sciences, Dhulikhel, Nepal

**Author notes:** **Corresponding Author** Prof. Dr. Rajeev Shrestha.

**Keywords:** iSeq100, Nepal, salmonella, whole genome sequencing

## Abstract

Bacterial whole genome sequencing helps to improve our understanding of epidemiology and pathogenesis of bacterial infections and allows comprehensive investigation on virulence, evolution and resistance mechanisms. Nepal, in recent times, has seen some increase in sequencing capabilities but faces numerous hurdles for optimum utilization. However, these hurdles can be alleviated with use of Illumina iSeq100. Therefore, this study aimed at performing whole genome sequencing of bacteria isolated utilizing the iSeq100 platform.

For this study, 6 banked *Salmonella enterica* serovar Typhi bacterial isolates were selected. These isolates were extracted for DNA, confirmed by qPCR and then, their libraries were prepared. The libaries were checked and loaded in Illumina iSeq100 at loading concentration of 200pM. The consensus was generated from the raw genomic data by reference-based assembly, mapping onto S. Typhi CT18. These consensus genomes and coverage parameters were compared to data from using HiSeq and NextSeq. The raw reads were also evaluated using pathogenwatch (v22.3.8) to observe for genotype, mutations and resistance genes.

The coverage parameters (coverage width and depth) of the genomes from this study were compared to same genomes sequenced using Illumina HiSeq and NextSeq. The average coverage width (96.81%) and depth (63.75x) of genomes sequenced in iSeq100 were comparable to that of HiSeq (width: 98.72% and depth: 69.87x). When the genomes sequenced were compared, the genotypes detected, number of SNPs and genetic determinants of AMR genes were identical.

The data from bacterial whole genome sequencing using the Illumina iSeq100 is equally informative when compared to other high-end sequencers. Therefore, the primary goal of this study is to advocate for optimum utlisation of iSeq100 while still ensuring a high standard of quality. This optimum utilization would create capacity to fill critical surveillance gaps.

## BACKGROUND

As part of most infection and outbreak management protocols, several phenotypic and molecular methods for pathogen characterization are conventionally utilized to monitor and control the spread of infections. [1] However, these conventional control approaches are slow and resource intensive while failing to distinguish between closely related strain, resistance and virulence factors, mostly due to limited genomic resolution and require multiple assays. [2-5] Therefore, for obtaining comprehensive information on phylogeny and improve outbreak investigations, full genomic information are needed. [2]

Bacterial whole genome sequencing (WGS) is one of the most promising approaches of this development, which helps to improve our understanding of epidemiology and pathogenesis of bacterial infections.[6] This approach due to its comprehensiveness on pathogen biology, mutations, drug resistance, transmission, evolution, community profiling, clinical metagenomics and pathogen discovery are already transforming the research landscape in microbiology.[7] This is evident by the large number of whole genomes stored in public repositories. [8,9] These approaches have known to be used in public health surveillance and control for several bacterial infections due to *Escherichia coli, Campylobacter jejuni, Legionella pneumophila and Mycobacterium tuberculosis*, outbreaks and monitor the source of healthcare associated infections. [10-14] Furthermore, continuous development in high throughput sequencing have pushed current clinical microbiology field due to its vast potential in identification of infectious agents, detection of pathogenicity, antimicrobial resistance, evoluation and epidemiological surveillance. [10,15-17] For instance, identification of single nucleotide polymorphisms (SNPs) can differentiate the evolution, with low frequency of SNPs indicating bacteria are genetically similar and recently originated from the same source. [2] Further, significance of WGS has been more evident by its use in COVID-19 pandemic. [18,19]

Although, WGS approach can comprehend all genomic information, it is only being utlilised in niches, because clinicians and researchers have shown reluctance, due higher costs, data interpretation and burdensome process of early sequencing technologies. [1,20,21] Further, some sequencing instruments have high operational costs, require large multiplexing for effective and cost continuous sequencing. [22] Nevertheless, the encouraging prospects are recent advancements in sequencing technologies (for instance, Sequencing by Synthesis from Illumina) and investigation tools have made the platform to have high throughput, increase output, decrease analysis time and reduced cost. [23,24]

Nepal, recently, has also seen some increase in sequencing potential, with some sites with next generation sequencing capabilities in Illumina, Nanopore and Thermo Fisher platforms. [25,26] However, not all the sequencers are at full potential due to technical sophistication, supply chain issue, limitation in capacity and funding and as a result, in most studies, samples are transported outside. [27] Nonetheless, these barriers of cost and technical complexity can be alleviated with optimum utilization Illumina iSeq 100, which is an inexpensive benchtop next-generation sequencer that minimizes the up-front instrument costs while maximizing simplicity of use and capability in obtaining bacterial whole genome, in country. [28,29]

Therefore, this study aimed at performing Whole Genome Sequencing (WGS) of bacteria isolated from Dhulikhel Hospital Kathmandu University Hospital, utilizing the Illumina iSeq100 platform.

## METHODS

### Bacterial Isolates Selection

For this study, 6 banked *Salmonella enterica* serovar Typhi bacterial isolates were selected randomly from Surveillance for Enteric Fever in Asia Project (SEAP). These isolates were previously sequencing in Illumina HiSeq and NextSeq platforms. These glycerol stock isolates were subcultured on MacConkey Agar (Oxoid, Cat: CM0007) and reconfirmed biochemically and serologically. An average of 30 isolated colonies were picked for each isolate.

### Processing, DNA Extraction and qPCR

The selected colonies were resuspended in 800ul of sterile normal saline, vortexed and centrifuged for 10 minutes at 8000rpm, to obtain cell pellets. The supernatant was discarded and then the pellets were resuspended in 200ul sterile 1XPBS. The pellets were subsequently revotexed and 100ul of the resuspension was taken for extraction.

The extraction was done as per manufacturer manual, using Qiagen DNeasy Blood and Tissue Kit. (Qiagen, Cat: 69504), with modification in elution. The final elution was done in 50ul, which was used for re-elution to increase the yield.

The S. Typhi isolated were re-confirmed by qPCR targeting Ty21a gene, using forward primer: 5’-CGCGAAGTCAGAGTCGACATAG-3’, reverse primer 5’-AAGACCTCAACGCCGATCAC-3’ and probe [6-FAM] CATTTGTTCTGGAGCAGGCTGACGG [BHQ1a-Q]. The assays were run in BioRad CFX 96 Dx qPCR machine with reaction mix that included: 10ul of 2X master mix (Quantabio Perfecta, Cat: 95113-012), 0.8ul of forward primer (400nM), 0.8 ul of reverse primer (400nM), 0.4ul of probe (200nM), 4ul of nuclease free water and 4ul of DNA template. The samples will Ct values <20.00 were selected for library preparation.

### Library Preparation and Whole Genome Sequencing

The concentration of the samples was checked by Qubit Fluorometer 4 (Invitrogen, Cat: Q33226) using dsDNA HS Kit (Invitrogen, Cat: Q32854). All the samples were diluted to have input amount of 75ng. The libraries were subsequently prepared using NEBNext Ultra II FS DNA Library Preparation Kit (New England Biolabs, Cat: 7805L). The libraries were quality checked for library size and concentration using Agilent Tapestation 4150 using D5000 HS Assay Kit (Agilent Technologies, Cat: 5067-5593). The average library size was 222.5bp. The isolates were loaded in two batches of two and four isolates at loading concentration of 200pM. The sequencing was done using pair ended barcoding primers at 2×146bp.

### Assembly, Consensus and Bioinformatical Investigation

The raw genomic data were trimmed for adapters (fastp), assembled (bowtie2, samtools), consensus generated, and coverage was calculated (bamCoverage, samtools depth, awk) and viewed in Integrated Genome Browser. The consensus was built using reference-based assembly and mapping on reference S. Typhi CT18. (Figure 1)

**Figure 1.**
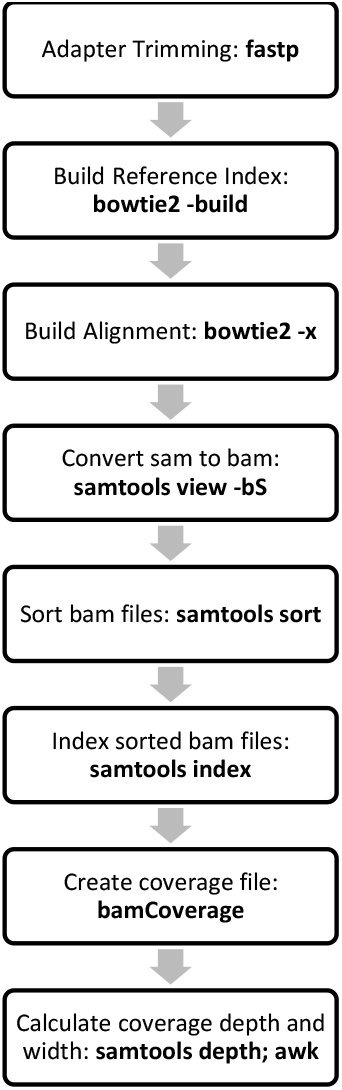
Bioinformatics workflow for generating and evaluating consensus genome

These consensus genomes and coverage parameters were compared to genomic data of same isolates previously sequenced using Illumina HiSeq (S3 to S6) and NextSeq (S1 and S2). These sequenced genomic data were downloaded from European Nucleotide Archive (ENA).

The raw reads were evaluated using Pathogenwatch (v22.3.8) to observe for genotype detected, number of Single Nucleotide Polymorphisms (SNPs), antimicrobial resistance determinants. Pathogenwatch is a platform which facilitates rapid identification of genomic markers of antimicrobial resistance (AMR) and includes latest analytics for typing along with epidemiological contextualization with public genomic data. [21]

### Data Availability

The genomic data, from Illumina iSeq100, can be obtained from National Center for Biotechnology Information’s BioProject PRJNA1110744 (Accession: SRX24530188 to SRX24530193).

## RESULTS

### Extraction and qPCR

The S. Typhi genomic DNA when evaluated for Ty21a gene, had average Ct value of 15.14, while the concentrations were variable, ranging from 1.58ng/ul to 20.6ng/ul.

### Whole Genome Sequencing

After the reference-based assembly, the Salmonella genomes were mapped over the S. Typhi CT18 strain in Integrated Genome Browser, with gaps observed in from ∼1,033,000bp to 1,047,000bp and ∼1,908,000bp to ∼1,933,000bp and ∼3,053,000bp to ∼3,059000bp. (Figure 2)

**Figure 2.**
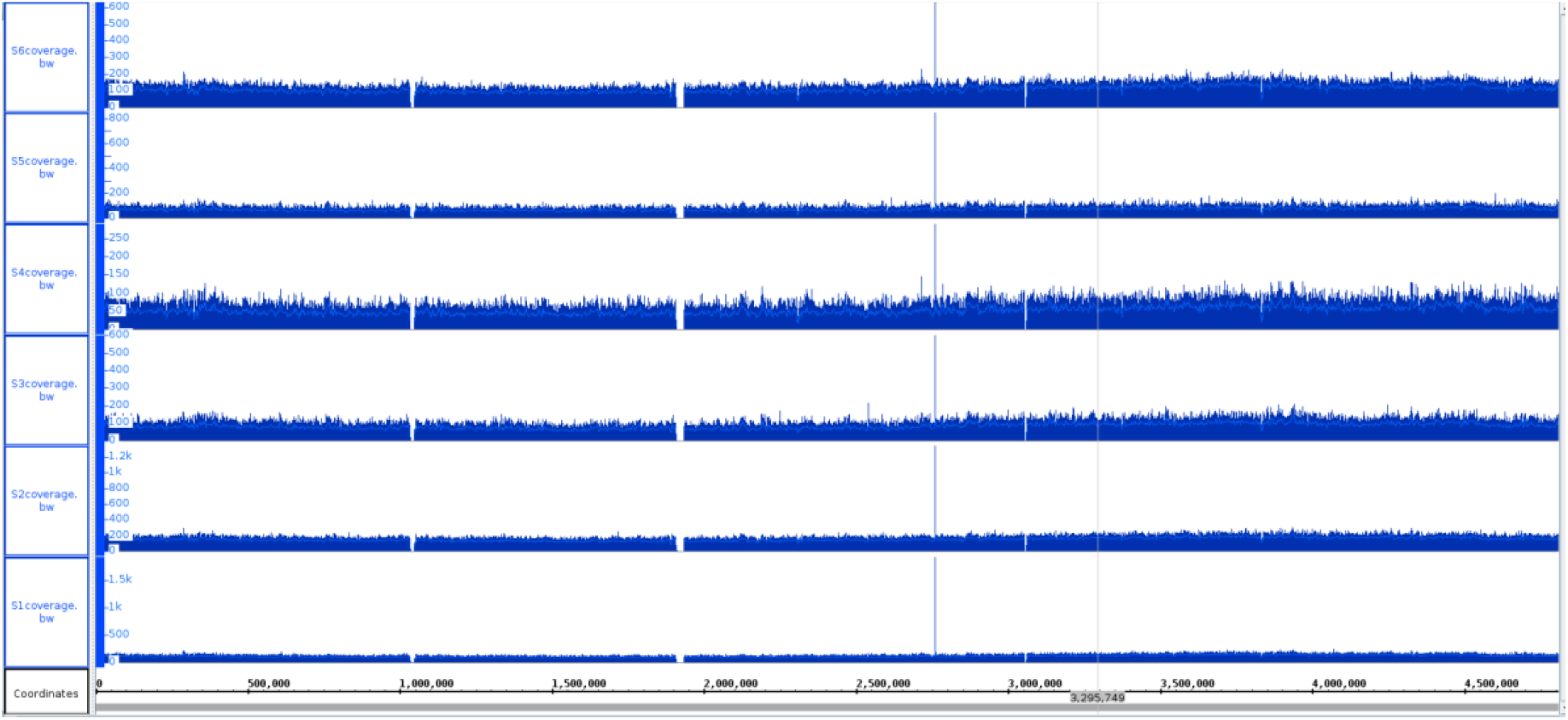
Coverage Visualization of S. Typhi genomes (S1 to S6) against S. Typhi CT18 strain reference genome, from Integrated Genome Browser

### Coverage Parameters

The coverage parameters (coverage width and depth) of the genomes from this study were compared to same genomes sequenced using Illumina HiSeq and NextSeq. The average coverage width (96.81%) and depth (63.75x) of genomes sequenced in iSeq100 were comparable to that of HiSeq and NextSeq (width: 98.72% and depth: 69.87x).

**Table 1:** Comparison of coverage depth and width between Illumina HiSeq and Illumina iSeq100.

|  | Illumina iSeq100 |  | Illumina HiSeq/NextSeq |  |
| --- | --- | --- | --- | --- |
| Sample | Average Coverage Depth | Coverage Width (%) at 20x | Average Coverage Depth | Coverage Width (%) at 20x |
| S1 | 72.23x | 98.91 | 50.43x | 98.90 |
| S2 | 101.91x | 98.93 | 92.77x | 99.97 |
| S3 | 52.40x | 98.41 | 65.47x | 98.92 |
| S4 | 34.23x | 89.04 | 88.49x | 98.94 |
| S5 | 46.67x | 97.00 | 46.81x | 97.05 |
| S6 | 75.02x | 98.58 | 75.29x | 98.58 |

### Comparison of Genotype, AMR genes and SNPs

When the genomes sequenced from Illumina iSeq100 and HiSeq/NextSeq were compared, the genotypes detected, number of SNPs and genetic determinants of AMR genes were identical.

**Table 2:**
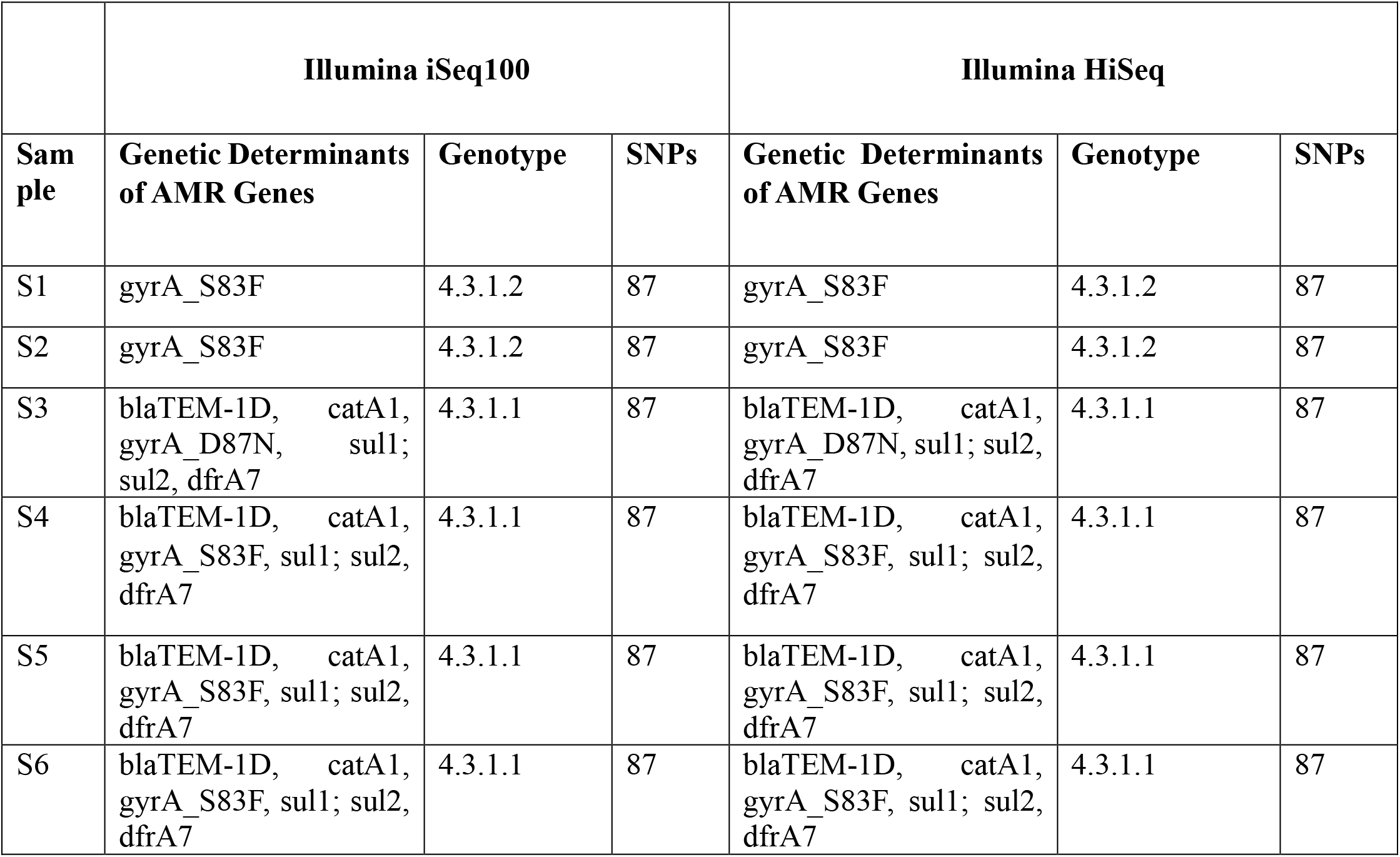
Comparison of genetic determinants, genotype and SNPs among genomes, evaluated by Pathogenwatch scheme (v22.3.8) for *Salmonella enterica* serovar Typhi (*S*. Typhi)

### Cost Evaluation

The sequencing was performed in two batches (two samples in first batch and four in second batch). When the collective cost of human resource, molecular and sequencing reagents and overhead was considered, the cost of sequencing bacterial genome in Illumina iSeq100 would be ∼$158 per gb when 4 samples are pooled and ∼$312 per gb when 2 samples are pooled.

## DISCUSSION

Despite the potential, widespread adoption of WGS has been hindered by various challenges, including high costs, time-consuming protocols, and data interpretation complexities. [30] The iSeq100 represents a significant advancement in sequencing technology, offering a cost-effective and user-friendly solution that overcomes many barriers associated with traditional NGS platforms.[29] This platform has been known to be more useful when used with amplicon-based sequencing wet-lab approach. [31-33] Therefore, this study addresses these hurdles by using a low-cost next generation sequencer, Illumina iSeq100 for WGS of *Salmonella Typhi*.

### Whole Genome Sequencing and Coverage Parameters

The coverage parameters of our approach were high (63.75x) and comparable to results from Illumina HiSeq and NextSeq (69.87x), which we agree could depend on the pooling. Though the threshold on the coverage depth depends upon the target pathogen, studies have estimated that that 40X are required for high-quality detection of virulence genes, while 21 bp reads at 4X coverage are sufficient for target gene detection and coverage width should preferably be between 50x – 100x. [34-36] In addition, in another study, for antibiotic resistance gene, at the read level, is considered present with 20x-30x coverage depth. [37,38] A study that utilized iSeq100 for WGS of bacterial isolates observed coverage depth of 17x - 149x with average of 59x, which is slightly lower than our observation. [28]

Similarly, high coverage width of at least 90% is recommended, which is well below the observed average coverage width of 96.81% of this study. [39] This high coverage width is required for optimum quality data during bioinformatics analysis, as this threshold helps achieve correct gene identification. [38]

The genome gaps were observed in the regions of ∼1,033,000bp to 1,047,000bp and ∼1,908,000bp to ∼1,933,000bp and ∼3,053,000bp to ∼3,059,000bp, which correspond to STY_RS04890 gene (DUF2213 domain-containing protein), WP_001197078.1 (baseplate J/gp47 family protein), STY_RS09550 (phage minor head protein), STY_RS09735 (tyrosine-type recombinase/integrase), STY_RS15100 (DUF1460 domain-containing protein) and STY_RS15130 (site-specific integrase). [40] These gaps could be a result of genomic rearrangements, horizontal gene transfer as well as sequencing and alignment errors and presence of low complexity regions. [41]

### Pooling and cost calculation

The pooling of isolates with varying genome size results in unequal sample coverage even with equimolar pooling, while some protocols do not mandatorily require equimolar pooling. [28,42] Further, pooling also depends on the complexity of the genome. For instance, though Escheria coli and Salmonella enterica have similar genome size, E. coli is more complex and has variable accessory genome. Therefore, comparatively larger number of S. enterica genomes can be pooled. [42] However, the cost per run also depends on pooling. The number of samples pooled would be inversely proportional to cartridge/flow cell requirement, while the amount of library preparation reagents remains the same. The average cost of run per gb is $60.65 for HiSeq (using HiSeq SBS Kit V4 kit - 250 cycles PE 2×125bp, which has now been discontinued), while for iSeq would be ∼$130 per gb when 4 samples are pooled and ∼$87 when 6 samples are pooled. [29,43] The cost, at our site, is slightly higher (∼$158 per gb when 4 samples are pooled), possibly because cost of sequencing is dependent on region and respective supply chain logistics. [44] Unfortunately, we cannot confirm the number of samples pooled for sequencing in HiSeq and NextSeq, to make more informed comparisons. During the discussion of low cost NGS, Oxford Nanopore provides great argument. The flongle, which gives output upto 2Gb, costs ∼$90, while iSeq100 cartridge/flowcell costs ∼$630 with 1.2Gb output. [42] However, the shorter shelf life of flow cell and lower accuracy of individuals reads are counter intuitive.

### Comparing genotypes and genomic determinants of AMR genes

In this study, both genotypes (4.3.1.1 and 4.3.1.2) belonging to lineage I and II, respectively, of H58 strain, were detected. Both strains were first detected in a pediatric study conducted in Kathmandu.[45] Furthermore, the point mutations (S83F and D87N) were observed in quinolone resistance determining region of gyrA gene, which is associated with fluoroquinolone resistance. [46-48] Similarly, mutations were observed in genes conferring resistance to ampicillin (blaTEM), chloramphenicol (catA1) and resistance to co-trimoxazole (sul) and resistance to trimethoprim (dfrA7). [49-52] In this study, the genotypes, genomic determinants and number of SNPs among the S. Typhi genomes were identical, as expected, in genomes sequenced from iSeq100 and HiSeq. This adds to the fact that the genomes sequenced have similar quality.

Our findings have demonstrated that iSeq platform is able to generate high-quality, accurate data for bacterial WGS, and could cost-effective addition to the genomic investigation capability. The genomes generated from iSeq have comparable coverage parameters to those obtained from more established commercial sequencing platforms like the Illumina HiSeq or NextSeq, which are not available, nor viable (due to its large number of sample requirements, reagents cost, annual maintenance cost among others) in Nepal. The findings of this study support premise that iSeq100 is well suited to small laboratories with low sample throughput, which is mostly the case in Nepal due to small molecular and genomic research ecosystem.

## CONCLUSION

The data from bacterial whole genome sequencing using the Illumina iSeq100 is equally informative when compared to other high-end sequencers. We have shown that Illumina iSeq, though primarily designed for microbial metagenomics, is able to perform whole genome sequencing of bacteria when combined with a modified wet-lab protocol. This approach will also provide high-quality genomes for other pathogens of human health importance, with some limitations on throughput. Thus, this study also advocates for optimum utlisation of iSeq100 while still ensuring a high standard of quality.

## Data Availability

The raw reads are available as an NCBI BioProject PRJNA1110744 (Accession: SRX24530188 to SRX24530193)

## IMPACT OF THE STUDY

Looking ahead, the findings of this study pave the way for broader applications of the Illumina iSeq100 platform in clinical microbiology, outbreak surveillance, and antimicrobial resistance monitoring in Nepal. Future research should focus on further optimizing sample preparation protocols, expanding sequencing capacity, and integrating bioinformatics tools to further enhance the utility and accessibility of WGS technologies in pathogen investigation. This approach will uplift the current national sequencing capabilities to full potential.

## DECLARATIONS

### Ethics approval and consent to participate

This study investigated banked bacterial isolates and did not contact human subjects or obtain data associated with them. The ethical approval was obtained from Institutional Review Committee at Kathmandu University School of Medical Sciences (ref: 115/24).

### Competing interests

The authors declare no competing interests.

### Funding

This study was funded and supported by Center for Infectious Disease Research and Surveillance, Dhulikhel Hospital Kathmandu University Hospital.

## Acknowledgements

We gratefully thank Jason R. Andrews and the SEAP team for providing accessions numbers (European Nucleotide Archive) for the *S. Typhi* isolates. We are also thankful to all the research staffs at Center for Infectious Disease Research and Surveillance, Dhulikhel Hospital Kathmandu University Hospital.

## Notes

### Competing Interest Statement

The authors have declared no competing interest.

### Summary of Updates

This revised version includes Figure 1, which was missing during pdf conversion, in previous version.

